# Artificial Intelligence-Enabled, Fully Automated Detection of Cardiac Amyloidosis Using Electrocardiograms and Echocardiograms

**DOI:** 10.1101/2020.07.02.20141028

**Authors:** Shinichi Goto, Keitaro Mahara, Lauren Beussink-Nelson, Hidehiko Ikura, Yoshinori Katsumata, Jin Endo, Hanna K. Gaggin, Sanjiv J. Shah, Yuji Itabashi, Calum A MacRae, Rahul C Deo

**Author notes:** **To whom correspondence should be addressed:** Rahul C Deo, MD, PhD, One Brave Idea / Division of Cardiovascular Medicine, Department of Medicine, Brigham and Women’s Hospital, Boston, MA, USA.

## Abstract

Although individually uncommon, rare diseases collectively affect over 350 million patients worldwide and are increasingly the target of therapeutic development efforts^1^. Unfortunately, the pursuit and use of such therapies have been hindered by a common challenge: patients with specific rare diseases are difficult to identify, especially if the conditions resemble more prevalent disorders. Cardiac amyloidosis is one such rare disease, which is characterized by deposition of misfolded proteins within the heart muscle resulting in heart failure and death^2–4^. In recent years, specific therapies have emerged for cardiac amyloidosis ^5,6^ and several more are under investigation^7–9^, but because cardiac amyloidosis is mistaken for common forms of heart failure, it is typically diagnosed late in its course. As a possible solution, artificial intelligence methods could enable automated detection of rare diseases, but model performance must address low disease prevalence. Here we present an automated multi-modality pipeline for cardiac amyloidosis detection using two neural-network models; one using electrocardiograms (ECG) and the second using echocardiographic videos as input. These models were trained and validated on 3 and 5 academic medical centers (AMC), respectively, in the United States and Japan. Both models had excellent accuracy for detecting cardiac amyloidosis with C-statistics of 0.85-0.92 and 0.91-1.00 for the ECG and echocardiography models, respectively, with the latter outperforming expert diagnosis. Simulating deployment on 13,906 and 7,775 patients with ECG-echocardiography paired data for AMC2 and AMC3 indicated a positive predictive value (PPV) for the ECG model of 4% and 3% at 61% and 54% recall, respectively. Pre-screening with ECG enhanced the echocardiography model performance from PPV 23% and 20% to PPV 58% and 53% at 64% recall, respectively for AMC2 and AMC3. In conclusion, we have developed a robust pipeline to augment detection of cardiac amyloidosis, which should serve as a generalizable strategy for other rare and intermediate frequency cardiac diseases with established or emerging therapies.

## Introduction

Cardiac amyloidosis arises from deposition of misfolded proteins in the heart muscle, which results in a restrictive-type cardiomyopathy, and commonly progresses to heart failure, conduction system disease, and cardiac death. Cardiac amyloidosis is subclassified based on the specific protein involved, with the major subtypes being transthyretin amyloidosis (ATTR cardiac amyloidosis), caused by misfolding of the transthyretin protein, and light chain amyloidosis (AL cardiac amyloidosis), caused by accumulation of immunoglobulin light chains^10^. Cardiac amyloidosis was previously believed to be rare but recent reports have suggested that the disease is largely underdiagnosed^11–15^. The imperative to identifying patients has dramatically increased with the advent of therapies for specific forms of cardiac amyloidosis^5–9^.

The clinical manifestations of cardiac amyloidosis – including conduction system disease, vitreous opacity, carpal tunnel syndrome, orthostatic hypotension, polyneuropathy, spinal stenosis, kidney dysfunction, atrial fibrillation, heart failure – are also commonplace in aging, thus making detection challenging. These signs and symptoms are distributed across multiple organs and tissues (and thus medical disciplines), and the probabilistic weighting of so many different features is forbidding, even in the unlikely event that all of the relevant exam findings, medical history details and diagnostic test results were available to a given practitioner. Furthermore, definitive diagnostic tests for cardiac amyloidosis – which include tissue biopsy and some forms of radionuclide scintigraphy – are costly and have associated risk, and thus are not plausible as screening approaches^2^.

Cardiac amyloidosis nonetheless has predictive features captured by less expensive and more widely available diagnostic modalities such as electrocardiography^16–19^ (ECG) and echocardiography^20–23^, but the features themselves are not highly specific and thus often missed. In addition, some of the recently highlighted echocardiographic features require providers to master specialized software packages^24^, which are time-consuming to use and thus tend to be employed in practice only after the disease is suspected. A truly generalizable detection strategy should require no specialized acquisition or processing and instead should use only widely available input data. However, the low existing prevalence of the disease places high demands on model performance to reduce the rate of costly false positives, something that has not been achieved to date. Here, we sought to develop a human-interpretation-free machine learning pipeline to detect cardiac amyloidosis using a combination of ECG and echocardiography.

## Results

### An ECG model detects cardiac amyloidosis effectively across multiple institutions

Electrocardiography is the most widely available cardiac diagnostic test and is frequently performed in primary care settings at low cost. Since many of the initial manifestations of cardiac amyloidosis are likely to result in presentation to a primary care physician, we sought first to develop a model based solely on ECG. We constructed ECG-derivation, ECG-validation and ECG-test groups from Brigham and Women’s Hospital (BWH) consisting of 5495, 2247 and 3191 ECG studies respectively (Figure S3, Methods). We tested the model’s performance using data from a held-out partition of the BWH data, as well as distinct cohorts from Massachusetts General Hospital (MGH) and University of California San Francisco (UCSF), which consisted of 842 and 1103 studies, respectively (Table1, Table 2). The composition of AL amyloidosis varied from 32.7 % to 58.5 % within these groups.

**Table 1.** Study-level demographic information (ECG cohort)

|  | BWH |  | MGH |  | UCSF |  |
| --- | --- | --- | --- | --- | --- | --- |
|  | Case | Control | Case | Control | Case | Control |
| Number studies | 2,249 | 8,684 | 405 | 437 | 372 | 731 |
| Age, years ± SD | 69.9 ± 10.4 | 62.3 ± 13.2 | 72.9 ± 9.0 | 73.8 ± 8.8 | 67.7 ± 12.9 | 67.5 ± 11.7 |
| Age Groups |  |  |  |  |  |  |
| $\leq 30$, n (%) | 2 (0.1) | 97 (1.1) | 1 (0.2) | 1 (0.2) | 2 (0.5) | 0 (0.0) |
| 30-50, n (%) | 78 (3.5) | 1,370 (15.8) | 7 (1.7) | 6 (1.4) | 36 (9.7) | 69 (9.4) |
| 50-70, n (%) | 901 (40.1) | 4,548 (52.4) | 143 (35.3) | 135 (30.9) | 136 (36.6) | 278 (38.0) |
| 70-90, n (%) | 1,242 (55.2) | 2,606 (30.0) | 254 (62.7) | 295 (67.5) | 198 (53.2) | 384 (52.5) |
| $>90$, n (%) | 26 (1.2) | 63 (0.7) | 0 (0.0) | 0 (0.0) | 0 (0.0) | 0 (0.0) |
| HR, bpm ± SD | 76.4 ± 16.7 | 75.9 ± 18.5 | 78.6 ± 16.6 | 75.1 ± 19.8 | 79.6 ± 18.7 | 72.2 ± 16.3 |
| Sinus rhythm, n (%) | 1,736 (77.2) | 8,072 (93.0) | 283 (69.9) | 371 (84.9) | 365 (98.1) | 729 (99.7) |

HR: heart rate, BWH: Brigham and Women's Hospital, MGH: Massachusetts General Hospital, UCSF: University of California San Francisco.
N represents the number of studies.

**Table 2.**
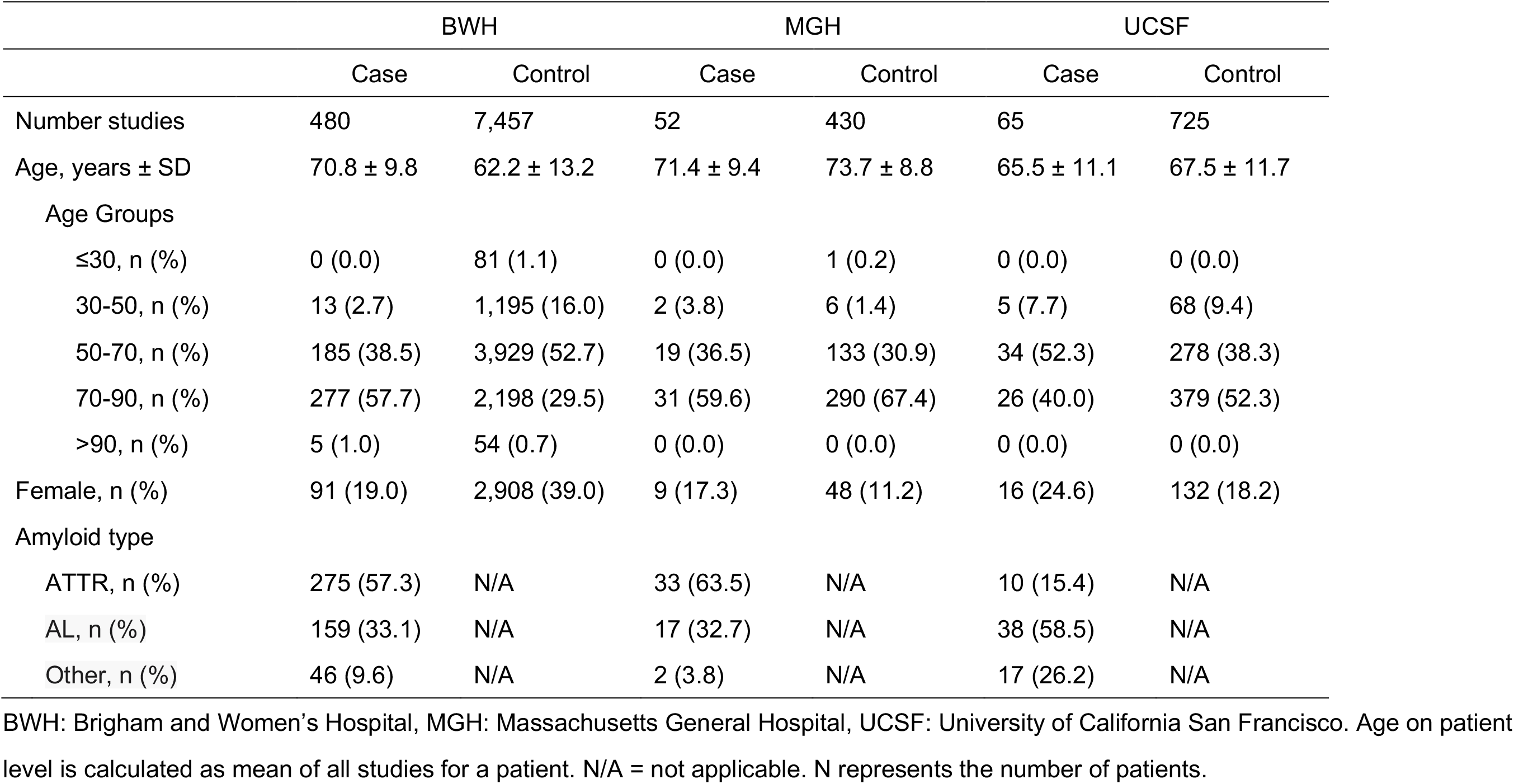
Patient-level demographic information (ECG cohort)

|  | BWH |  | MGH |  | UCSF |  |
| --- | --- | --- | --- | --- | --- | --- |
|  | Case | Control | Case | Control | Case | Control |
| Number studies | 480 | 7,457 | 52 | 430 | 65 | 725 |
| Age, years ± SD | 70.8 ± 9.8 | 62.2 ± 13.2 | 71.4 ± 9.4 | 73.7 ± 8.8 | 65.5 ± 11.1 | 67.5 ± 11.7 |
| Age Groups |  |  |  |  |  |  |
| $\leq 30$, n (%) | 0 (0.0) | 81 (1.1) | 0 (0.0) | 1 (0.2) | 0 (0.0) | 0 (0.0) |
| 30-50, n (%) | 13 (2.7) | 1,195 (16.0) | 2 (3.8) | 6 (1.4) | 5 (7.7) | 68 (9.4) |
| 50-70, n (%) | 185 (38.5) | 3,929 (52.7) | 19 (36.5) | 133 (30.9) | 34 (52.3) | 278 (38.3) |
| 70-90, n (%) | 277 (57.7) | 2,198 (29.5) | 31 (59.6) | 290 (67.4) | 26 (40.0) | 379 (52.3) |
| $>90$, n (%) | 5 (1.0) | 54 (0.7) | 0 (0.0) | 0 (0.0) | 0 (0.0) | 0 (0.0) |
| Female, n (%) | 91 (19.0) | 2,908 (39.0) | 9 (17.3) | 48 (11.2) | 16 (24.6) | 132 (18.2) |
| Amyloid type |  |  |  |  |  |  |
| ATTR, n (%) | 275 (57.3) | N/A | 33 (63.5) | N/A | 10 (15.4) | N/A |
| AL, n (%) | 159 (33.1) | N/A | 17 (32.7) | N/A | 38 (58.5) | N/A |
| Other, n (%) | 46 (9.6) | N/A | 2 (3.8) | N/A | 17 (26.2) | N/A |

BWH: Brigham and Women's Hospital, MGH: Massachusetts General Hospital, UCSF: University of California San Francisco. Age on patient level is calculated as mean of all studies for a patient. N/A = not applicable. N represents the number of patients.

The ECG model showed good predictive accuracy as measured by area under the receiver operating test characteristic curve (ROC-AUC) of 0.91 (95% CI 0.90-0.93) on the ECG-test set of BWH and similar performance with ROC-AUC of 0.85 (0.82-0.87) on Massachusetts General Hospital (MGH) cohort and 0.86 (0.83-0.88) for the University of California San Francisco (UCSF) cohort (**Figure 1**). A sensitivity analysis to amyloidosis subtype demonstrated slightly better performance on ATTR amyloid with AUC of 0.94 (0.92-0.95), 0.87 (0.84-0.89), 0.97 (0.95-0.98) when compared to AL amyloid which showed AUC of 0.92 (0.89-0.94), 0.81 (0.76-0.86) and 0.78 (0.75-0.82) for BWH, MGH and the UCSF cohorts respectively (**Figure S1**).

**Figure 1.**
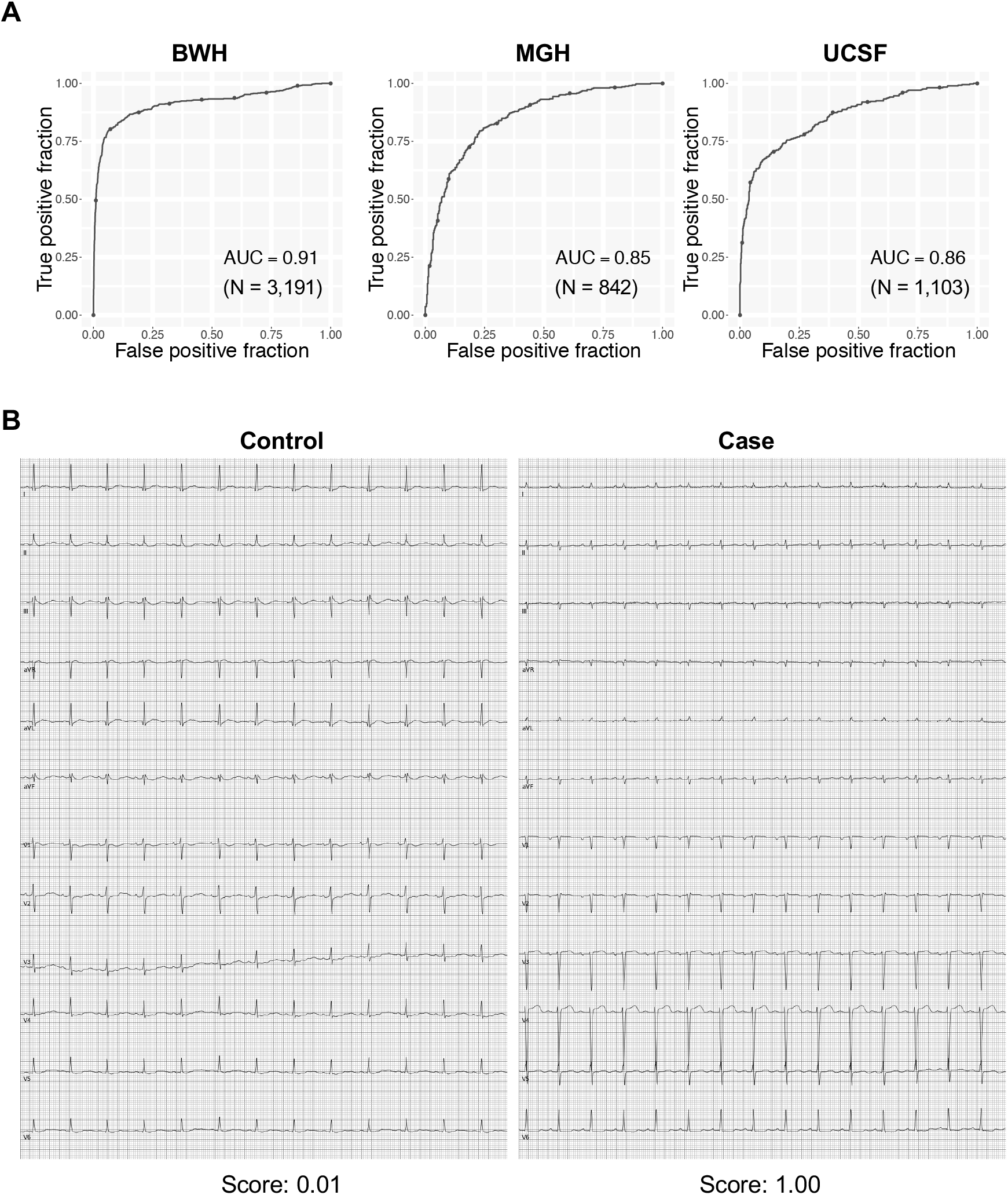
Performance of the cardiac amyloidosis ECG model. (A) ROC plots for detecting cardiac amyloidosis for each institution. The performance on a test dataset is shown for BWH. (B) Representative ECG for cases and controls. The score denotes the model output for the video. N is the numbers of studies.

### A video-based echocardiography model for cardiac amyloidosis has excellent performance for patients from 5 AMCs across 2 countries

Although the ECG-based models were encouraging, we anticipated they did not have the requisite performance characteristics for a low prevalence disease. We thus trained an echocardiogram video-based model, using only a single commonly acquired view, the apical 4-chamber view (A4C), which can be collected even with low-cost handheld ultrasound devices. The echocardiogram-derivation, echocardiogram-validation and echocardiogram-test group from BWH had 5257, 2136 and 3410 videos respectively. The external validation cohorts from MGH, UCSF, Northwestern University (NW) and Keio University Hospital (Keio) in Japan had 441, 369, 229 and 239 studies for 361, 350, 200, and 173 patients respectively (Tables 3 and 4).

**Table 3.** Study-level demographic information (Echocardiogram cohort)

|  | BWH |  | MGH |  | UCSF |  | NW |  | Keio |  |
| --- | --- | --- | --- | --- | --- | --- | --- | --- | --- | --- |
|  | Case | Control | Case | Control | Case | Control | Case | Control | Case | Control |
| Number studies | 1,486 | 2,213 | 110 | 331 | 46 | 323 | 103 | 126 | 118 | 121 |
| Age, years ± SD | 72.9 ± 8.8 | 70.0 ± 11.7 | 71.9 ± 9.7 | 70.2 ± 12.3 | 70.3 ± 11.6 | 67.0 ± 12.8 | 75.1 ± 7.4 | 74.7 ± 7.6 | 74.2 ± 8.2 | 73.8 ± 8.5 |
| Age Groups |  |  |  |  |  |  |  |  |  |  |
| ≤30, n (%) | 0 (0.0) | 8 (0.4) | 0 (0.0) | 0 (0.0) | 0 (0.0) | 0 (0.0) | 0 (0.0) | 0 (0.0) | 0 (0.0) | 0 (0.0) |
| 30-50, n (%) | 27 (1.8) | 111 (5.0) | 7 (6.4) | 36 (10.9) | 3 (6.5) | 37 (11.5) | 0 (0.0) | 0 (0.0) | 0 (0.0) | 0 (0.0) |
| 50-70, n (%) | 479 (32.2) | 940 (42.5) | 34 (30.9) | 108 (32.6) | 15 (32.6) | 137 (42.4) | 27 (26.2) | 39 (31.0) | 30 (25.4) | 34 (28.1) |
| 70-90, n (%) | 967 (65.1) | 1,118 (50.5) | 69 (62.7) | 181 (54.7) | 28 (60.9) | 149 (46.1) | 75 (72.8) | 87 (69.0) | 86 (72.9) | 83 (68.6) |
| >90, n (%) | 13 (0.9) | 36 (1.6) | 0 (0.0) | 6 (1.8) | 0 (0.0) | 0 (0.0) | 1 (1.0) | 0 (0.0) | 2 (1.7) | 4 (3.3) |
| HR, bpm ± SD | 74.7 ± 18.4 | 73.8 ± 20.5 | 79.4 ± 21.5 | 71.4 ± 16.1 | 78.0 ± 14.7 | 74.1 ± 21.9 | 74.7 ± 19.5 | 63.8 ± 9.9 | 71.8 ± 17.0 | 69.4 ± 15.4 |
| Manufacture |  |  |  |  |  |  |  |  |  |  |
| Philips, n (%) | 264 (17.8) | 1,655 (74.8) | 105 (95.5) | 313 (94.6) | 44 (97.5) | 312 (96.6) | 0 (0.0) | 0 (0.0) | 48 (40.7) | 38 (31.4) |
| GE, n (%) | 1,211 (81.5) | 553 (25.0) | 5 (4.5) | 18 (5.4) | 2 (4.3) | 11 (3.4) | 103 (100.0) | 126 (100.0) | 47 (39.8) | 57 (47.1) |
| Agilent, n (%) | 3 (0.2) | 0 (0.0) | 0 (0.0) | 0 (0.0) | 0 (0.0) | 0 (0.0) | 0 (0.0) | 0 (0.0) | 0 (0.0) | 0 (0.0) |
| Sono Site, n (%) | 1 (0.1) | 0 (0.0) | 0 (0.0) | 0 (0.0) | 0 (0.0) | 0 (0.0) | 0 (0.0) | 0 (0.0) | 0 (0.0) | 0 (0.0) |
| SIEMENS, n (%) | 6 (0.4) | 5 (0.2) | 0 (0.0) | 0 (0.0) | 0 (0.0) | 0 (0.0) | 0 (0.0) | 0 (0.0) | 0 (0.0) | 0 (0.0) |
| TOSHIBA, n (%) | 0 (0.0) | 0 (0.0) | 0 (0.0) | 0 (0.0) | 0 (0.0) | 0 (0.0) | 0 (0.0) | 0 (0.0) | 23 (19.5) | 26 (21.5) |

HR: heart rate, BWH: Brigham and Women's Hospital, MGH: Massachusetts General Hospital, UCSF: University of California San Francisco, NW:
North Western University, Keio: Keio University Hospital. N represents the number of studies.

**Table 4.** Patient-level demographic information (Echocardiogram cohort)

|  | BWH |  | MGH |  | UCSF |  | NW |  | Keio |  |
| --- | --- | --- | --- | --- | --- | --- | --- | --- | --- | --- |
|  | Case | Control | Case | Control | Case | Control | Case | Control | Case | Control |
| Number studies | 421 | 2,069 | 41 | 320 | 32 | 318 | 74 | 126 | 52 | 121 |
| Age, years ± SD | 72.7 ± 9.4 | 69.7 ± 11.7 | 71.0 ± 10.1 | 70.3 ± 12.4 | 69.2 ± 12.0 | 67.0 ± 12.8 | 75.6 ± 7.3 | 74.7 ± 7.6 | 74.2 ± 8.3 | 73.8 ± 8.5 |
| Age Groups |  |  |  |  |  |  |  |  |  |  |
| ≤30, n (%) | 0 (0.0) | 8 (0.4) | 0 (0.0) | 0 (0.0) | 0 (0.0) | 0 (0.0) | 0 (0.0) | 0 (0.0) | 0 (0.0) | 0 (0.0) |
| 30-50, n (%) | 9 (2.1) | 108 (5.2) | 3 (7.3) | 36 (11.3) | 2 (6.2) | 36 (11.3) | 0 (0.0) | 0 (0.0) | 0 (0.0) | 0 (0.0) |
| 50-70, n (%) | 143 (34.0) | 898 (43.4) | 14 (34.1) | 101 (31.6) | 12 (37.5) | 136 (42.8) | 17 (23.0) | 39 (31.0) | 13 (25.0) | 34 (28.1) |
| 70-90, n (%) | 264 (62.7) | 1,024 (49.5) | 24 (58.5) | 177 (55.3) | 18 (56.2) | 146 (45.9) | 56 (75.7) | 87 (69.0) | 39 (75.0) | 83 (68.6) |
| >90, n (%) | 5 (1.2) | 31 (1.5) | 0 (0.0) | 6 (1.9) | 0 (0.0) | 0 (0.0) | 1 (1.4) | 0 (0.0) | 0 (0.0) | 4 (3.3) |
| Female, n (%) | 76 (18.1) | 711 (34.4) | 12 (29.2) | 133 (41.6) | 10 (31.3) | 71 (22.3) | 8 (10.8) | 14 (11.1) | 6 (11.5) | 16 (13.2) |
| Amyloid type |  |  |  |  |  |  |  |  |  |  |
| ATTR, n (%) | 287 (68.2) | N/A | 21 (51.2) | N/A | 9 (28.1) | N/A | 74 (100.0) | N/A | 45 (86.5) | N/A |
| AL, n (%) | 125 (29.7) | N/A | 19 (46.3) | N/A | 11 (34.4) | N/A | 0 (0.0) | N/A | 7 (13.5) | N/A |
| Other, n (%) | 9 (2.1) | N/A | 1 (2.4) | N/A | 12 (37.5) | N/A | 0 (0.0) | N/A | 0 (0.0) | N/A |

BWH: Brigham and Women's Hospital, MGH: Massachusetts General Hospital, UCSF: University of California San Francisco, NW: North Western University, Keio: Keio University Hospital. N represents the number of patients. Age is calculated as mean of all studies for a patient. N represents the number of patients.

**Table 5.** Demographic information for deployment simulation cohort

|  | MGH | UCSF |
| --- | --- | --- |
| Number patients | 13,906 | 7,794 |
| Age, years ± SD | 65.8 ± 16.1 | 60.8 ± 17.8 |
| Age Groups |  |  |
| $\leq 30$, n (%) | 470 (3.4) | 470 (6.0) |
| 30-50, n (%) | 1,753 (12.6) | 1,489 (19.1) |
| 50-70, n (%) | 5,500 (39.6) | 3,179 (50.8) |
| 70-90, n (%) | 5,704 (41.0) | 2,423 (31.1) |
| $> 90$, n (%) | 480 (3.5) | 233 (3.0) |
| Female, n (%) | 6,101 (43.9) |  |
| HR, bpm ± SD | 72.2 ± 17.7 | 74.5 ± 19.0 |
| Manufacture |  |  |
| Philips, n (%) | 13,239 (95.2) | 6,996 (89.8) |
| GE, n (%) | 667 (4.8) | 795 (10.2) |
| SIEMENS, n (%) | 0 (0.0) | 2 (0.0) |
| ACUSON, n (%) | 0 (0.0) | 1 (0.0) |

HR: heart rate, MGH: Massachusetts General Hospital. UCSF: University of California San Francisco

The echocardiography model showed excellent predictive accuracy, with ROC-AUC of 0.97 (0.96-0.98) on the BWH test dataset, and similar performances on external validation cohorts from 3 institutions of US and 1 from Japan with ROC-AUC of 0.91 (0.88-0.94) for MGH, 0.93 (0.88-0.97) for UCSF, 1.00 (1.00-1.00) for NW and 0.95 (0.91-0.97) for Keio (**Figure 2**). Analysis on cardiac amyloidosis subtypes showed superior model performance on ATTR amyloid with AUC of 0.98 (0.97-0.99), 0.95 (0.91-0.98), 0.98 (0.95-1.00), 1.00 (1.00-1.00) and 0.95 (0.91-0.97) for BWH, MGH, UCSF, NW, and Keio when compared to AL amyloid which had an AUC of 0.95 (0.93-0.97), 0.86 (0.81-0.92), 0.87 (0.77-0.96) and 0.92 (0.86-0.97) for BWH, MGH, UCSF and Keio (the NW dataset had no AL amyloid cases) (**Figure S2**).

**Figure 2.**
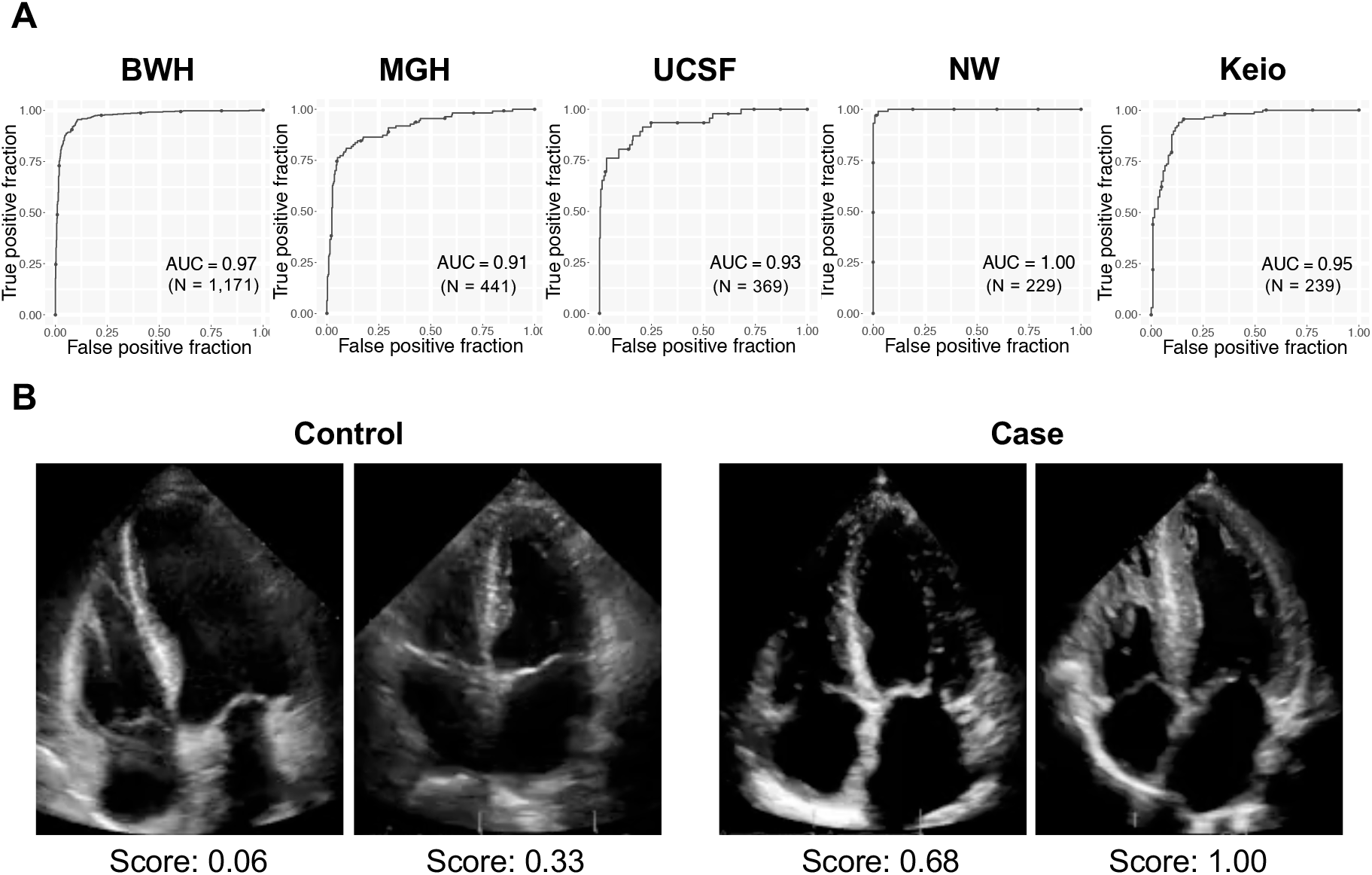
Performance of the cardiac amyloidosis echocardiogram model. (A) ROC plots for detecting cardiac amyloidosis for each institution. The performance on the test dataset is shown for BWH. (B) representative echo images for cases and controls. The score denotes the model output for the video. N is the numbers of studies.

### The cardiac amyloidosis echocardiography model outperforms interpretation by expert cardiologists

Two issues make detection of cardiac amyloidosis on echocardiograms particularly challenging for human readers: a lack of sufficiently specific features within the videos and the triggering required to screen for these features. Although the latter is difficult to address within existing clinical workflows (though completely solved by an automated system), we sought to evaluate the former by head-to-head comparison. We thus had two expert readers (KM, SG) attempt to diagnose cardiac amyloidosis using the test sets from 3 institutions: MGH, UCSF, and Keio (**Figure 3)**. In all cases, the model AUC outperformed the human readers (Figure 3), though for KM in UCSF, the result was within the 95% confidence interval. Overall, the model’s superior performance was more apparent for ATTR than AL amyloidosis.

**Figure 3.**
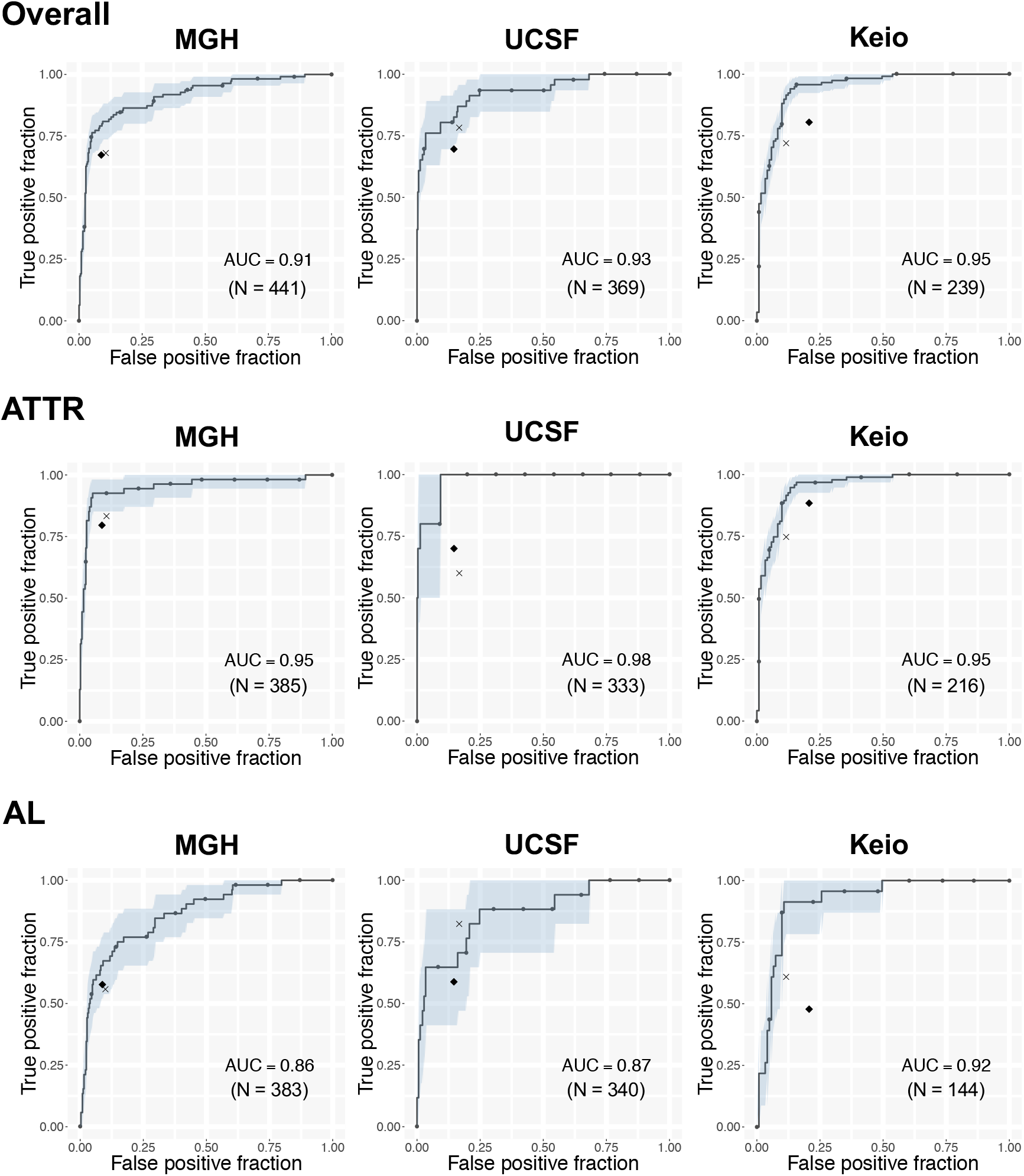
Comparison of the echocardiogram model with expert interpretation. ROC plots for detecting cardiac amyloidosis for each institution and amyloid type. The area in light blue represents the 95% CI for the true positive fraction for a given false positive fraction. The black diamond represents the performance of the general cardiologist interpretation and the *x* represents the performance of the echocardiography expert cardiologist for detecting cardiac amyloid. N is the numbers of studies.

### Stepwise approach using ECG and echocardiography models detects cardiac amyloidosis from a surveillance population

Within the MGH and UCSF cohorts, there were 13,906 patients and 7794 patients with ECG-echocardiogram pairs (within 2 years of one another, with the ECG preceding the echocardiogram) respectively. Based on the output of the echocardiogram model, we estimated the prevalence of cardiac amyloidosis in this group was 0.83% and 0.69%, which is in keeping with our estimates of cardiac amyloidosis prevalence within this population (see Methods). Using the echocardiography output as gold standard, the ECG model detected cardiac amyloidosis with PPV 3.8% with recall 61.2% in MGH and PPV 3.1% with recall 53.7% in UCSF at a cutoff 0.5 (**Figure 4A**). Using the AUROC curve to estimate a likelihood ratio and the above estimated prevalence numbers, the echocardiogram model alone detected cardiac amyloidosis with a PPV of 22.9% with recall 64.3% for MGH and PPV 19.6% with recall 63.9% for UCSF at a cutoff of 0.9 (**Figure 4B**). Assuming an updated prevalence after pre-screening using the ECG model, the PPV improved to 58.3% for MGH and 53.1% for UCSF with the same cutoff. The combined ECG-echocardiogram pipeline thus resulted in an overall recall of 39.4% and 34.3% for MGH and UCSF, respectively at a PPV of 64% (**Figure 4C)**. In comparison, for those same PPV values, the echocardiogram model alone could only achieve recall of 11.6% for MGH and recall of 11.4% for UCSF.

**Figure 4.**
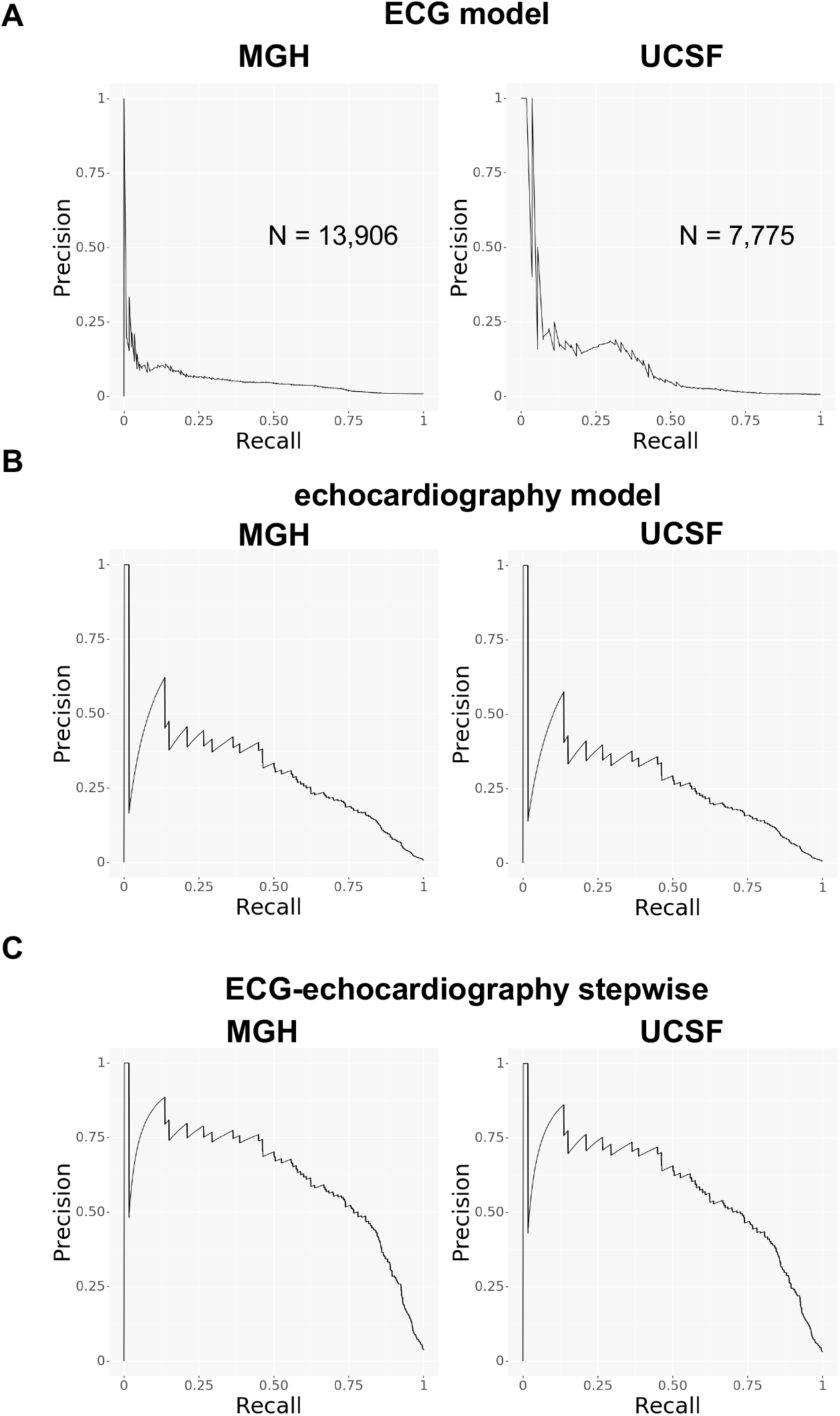
Screening performance of the models on surveillance populations. Precision recall curve plots for (A) the ECG model, (B) the echocardiogram model, and (C) the echocardiogram model after ECG pre-screening for detection of cardiac amyloidosis in surveillance populations. N is the numbers of studies.

## Discussion

Cardiac amyloidosis is one member of a group of cardiovascular diseases, including hypertrophic cardiomyopathy and pulmonary arterial hypertension, that is potentially treatable but rare and therefore difficult to detect. The imperative to recognize patients with these and other rare diseases largely depends on availability of specific therapeutic options, but once these appear, it can be difficult to rapidly adapt prior workflows to ensure that patients are treated appropriately. Moreover, since patients are likely to present to non-experts with their initial symptoms, an operational challenge becomes how best to construct systems that facilitate detection even in such settings.

Although the impact of cardiac amyloidosis on ECG and echocardiography has been known for many decades, the features themselves in isolation have not been sufficiently specific or sensitive to be used as heuristics. For example, in one study of 40 cardiac amyloidosis patients, the characteristic low-voltage ECG pattern of cardiac amyloidosis was seen in only 18% of cardiac amyloidosis patients. One could in principle combine these with other non-cardiac features, but this places an increasing burden on the provider to seek such information, which often only occurs when a suspicion of the disease exists in the first place.

In contrast, the approach we have developed here has deliberately limited the need for any recognition by the provider and use inputs that can be acquired in primary care settings – whether by ECG or handheld echocardiography. To further enable effective deployment in such settings, these detection approaches should ideally be coupled with further facilitation of confirmatory diagnostic processes. In fact, our approach benefits from the fact that there is this second gate of confirmatory diagnostic testing: namely measurement of free light chains, scintigraphy scanning, and possibly tissue biopsy^3^. The ECG and echocardiography models thus represent a tunable detection tool, with cutpoints that can be selected based on population prevalence and costs and benefits (diagnostic, therapeutic, financial and otherwise) of downstream true and false positives (and negatives). The data collected through deployment can itself enable refinement of cutpoints, and potentially spur retraining of models to better match local conditions. Critically, in such a system involving a confirmatory step downstream of the AI detection output, model explainability is less of an issue, and one can focus on maximizing model performance.

Medicine has historically reserved screening for widely prevalent diseases such as breast and colon cancer, in part because of the larger number of individuals who may benefit, and also because of the anticipated higher PPV of any diagnostic algorithms. However, given the collective scope of rare diseases^1^, the possibility of developing highly specific models to recognize them (whether by genetics or imaging), and the increasing number of therapies being developed to target them, it will be informative to establish whether a similar paradigm can be developed for other underdiagnosed conditions.

## Methods

### Patient selection procedure for ECG and echocardiography models

For all institutions, prospective cardiac amyloidosis patients were first identified based on diagnostic codes and/or echocardiography reports and then manually confirmed by chart review. Specifically, patients with ATTR cardiac amyloidosis were required to have 1) confirmation of amyloid disease by tissue biopsy, nuclear medicine scan, cardiac magnetic resonance imaging, or genetic testing (transthyretin variant) or 2) strong suspicion based on characteristic features of the echocardiogram (these tended to be patients from an era prior to widespread scintigraphy testing). For AL amyloid, biopsy confirmation was required as well as some evidence of cardiac involvement, whether by cardiac magnetic resonance or echocardiography. For both models, they were matched based on age and sex to patients who underwent ECG or echocardiography at the same institution but did not have cardiac amyloidosis. For the ECG cohorts, ECGs with pacing spikes were excluded.

The ECG model was trained with data from Brigham and Women’s Hospital (BWH) and was externally validated with the data from 2 different institutions from US: Massachusetts General Hospital (MGH) and University of California San Francisco (UCSF). The patients from BWH were randomly split into 3 groups (ECG-Derivation, ECG-validation and ECG-Test cohort) in a 5:2:3 ratio to be used for model training **(Figure S3)**. Patients who had ECGs at both BWH and MGH were identified and was allocated to the ECG-Test cohort to avoid overfitting.

The echocardiography model was trained with data from Brigham and Women’s Hospital (BWH) and was externally validated with the data from 4 different institutions from US and Japan: Massachusetts General Hospital (MGH), University of California San Francisco (UCSF), North-Western University (NW) and Keio University Hospital (Keio) **(Figure S4, S5, S6, S7 and S8)**. To make the model robust to intracardiac leads, An additional 256 patients with pacemaker or ICD lead and without cardiac amyloidosis was identified and added to the control for the BWH dataset. The patients from BWH were randomly split into 3 groups (echocardiogram-derivation, echocardiogram-validation and echocardiogram-test cohort) in a 5:2:3 ratio to be used for model training. Patients who had an echocardiography study at both BWH and MGH were identified and was allocated to the echocardiogram-test cohort to avoid overoptimistic estimation of model performance on the MGH test set.

### ECG model architecture and training

The ECG model was constructed as a 2D-CNN based model. It consisted of a layer of 2D-CNN followed by 18 layers of multi-2D-CNN-module, which was constructed by 3 parallel multilayer CNNs concatenated at the end of the module (schematic shown in **Figure S9**, code is included ECGModel.py). The model had 49,823,214 parameters total and 49,744,020 were trainable. The model was trained using data from ECG-Derivation cohort from BWH. ECGs were labeled as case=1 or control=0 and the model was trained to minimize the binary cross entropy between model prediction and the label using RMSprop optimizer with initial learning rate of 0.0001. The model was trained for 150 epochs. At the end of each epoch, ROC-AUC on the ECG-validation cohort was calculated. The final model was chosen as the model with highest ROC-AUC on the validation cohort across all 150 epochs.

### Echocardiogram model architecture and training

Given that echocardiograms are videos, which are time-series of multiple frames, we constructed a 3D-CNN based model treating temporal axis as the 3^rd^ axis rather than taking a frame-by-frame approach as done previously^20^, to maximize the ability of the model to use dynamic features in disease detection. It consisted of 3 layers of 3D-CNN followed by 12 layers of Multi-3D-CNN-module, which was constructed by 3 parallel multilayer 3D-CNNs and a max pooling operation concatenated at the end of the module (schematic shown in **Figure S10**, code is included as EchoModel.py). The scales of the video (in cm/pixel) was input into the fully connected layer. The model had 28,341,385 parameters total and 28,298,105 were trainable. The model was trained using data from echocardiogram-derivation cohort from BWH. The echocardiography videos were labeled as case=1 or control=0 on the study level and was trained to minimize the binary cross entropy between model prediction and the label using RMSprop optimizer with initial learning rate of 0.0001. The model was trained for 50 epochs. At the end of each epoch, ROC-AUC on ECG-validation cohort was calculated. The final model was chosen as the model with highest ROC-AUC on validation cohort across all 50 epochs.

### Echocardiography model comparison with expert cardiologist interpretation

The performance of the echocardiography model to detect cardiac amyloidosis was compared with two expert cardiologists (SG: general cardiologist and MK: National Board-certified expert in Adult Comprehensive Echocardiography). The comparison was performed at the study level rather than individual video level. While the CNN model diagnostic output was based on only apical 4 chamber views, the experts had access to all the videos in each echocardiogram study to diagnose cardiac amyloidosis. The experts were blinded to model output. The experts labeled each study as cardiac amyloidosis positive or negative for 3 external validation datasets from MGH, UCSF and Keio. Sensitivity and specificity were calculated and compared with the ROC curve of the model. A subtype analysis on ATTR and AL amyloidosis was also performed.

### Estimating positive predictive value of ECG, echocardiogram, and combined ECG-echocardiogram models

We estimated prevalence for cardiac amyloidosis within the population of patients with echocardiograms as follows. From our internal data across two large AMCs, we have found that over the past 4 years, 20-25% of the ∼16,000-18,000 unique patients who obtain an echocardiogram have at least one encounter diagnosis for heart failure. Of those we anticipate 50% to have heart failure with preserved ejection fraction (HFpEF), or 10-12.5% of patients. The percentage of cardiac amyloidosis within HFpEF is unknown but recent studies suggest proportions of 13-20% in selected subsets^11–15^. Given that these represented enriched populations, we assumed a lower value of 5-7%, which corresponds to 0.5-0.9% of our total population. This value is in keeping with prevalence analysis using 916 successive echocardiograms from Keio University, which included 7 patients with known cardiac amyloidosis (0.76%).

To estimate PPV for our ECG model, we identified 13,906 and 7775 patients within our respective MGH and UCSF cohorts with an ECG followed by an echocardiogram within 2 years. (**Figure S11 and S12**). A single ECG-echocardiography study pair was selected for each patient that had the shortest time between ECG and echocardiography studies. We deployed the ECG and echocardiogram cardiac amyloidosis models on each study and defined the gold standard as individuals with an echocardiogram model score of at least 0.9, a threshold that resulted in prevalence values of 0.83% and 0.69% for MGH and UCSF, respectively. We assessed the ability of ECG model to detect cardiac amyloidosis using precision-recall curve plots.

To assess the PPV for the echocardiogram model, we estimated a likelihood ratio from the receiver operating characteristic curve^25^ across the combined test sets for BWH, MGH, UCSF, and Keio. At a threshold of 0.9, the likelihood ratio of the echocardiogram model was 35.4. Assuming the above cardiac amyloidosis prevalence of 0.83% and 0.69% for MGH and UCSF, respectively, we were able to estimate an institution-level PPV for the echocardiogram model. For the successive deployment of ECG and echocardiogram models, we updated the PPV based on the prevalence expected from using only studies that exceeded a cutpoint of 0.5 from the output of the ECG model.

### Statistical analysis

All the models were trained with *Keras 2.3.0* on a *Tensorflow 1.14.0* backend^29^. The ROC curves are plotted using the *ggplot2*^27^ package (R 3.6.1) and the AUC-ROC, sensitivity, specificities, and 95% confidence intervals (using 2000 bootstrap samples) were calculated using *pROC*^28^ package. The precision-recall plots were made using the *plotnine* package in Python 3.7. Continuous values are presented as mean ± standard deviation (SD) and categorical values are presented as numbers and percentages if not otherwise specified.

## Data Availability

The code of the model is provided as supplementary material attached to this manuscript.

## Supplementary Figure legends

**Figure S1.**
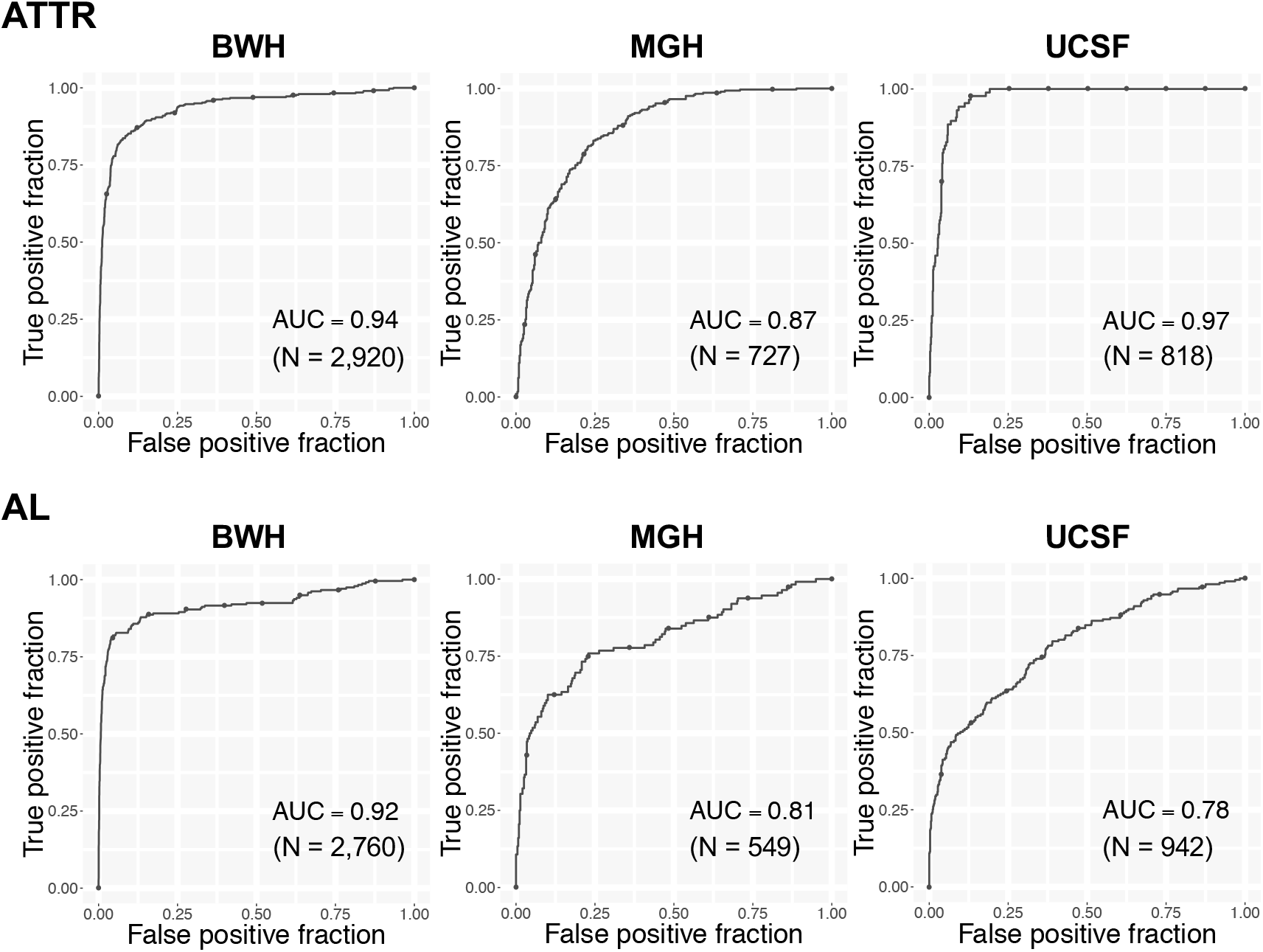
Performance of ECG model by amyloid subtype. ROC plots for detecting cardiac amyloidosis on amyloid subtype for each institution. The performance on a test dataset is shown for BWH. N is the numbers of studies

**Figure S2.**
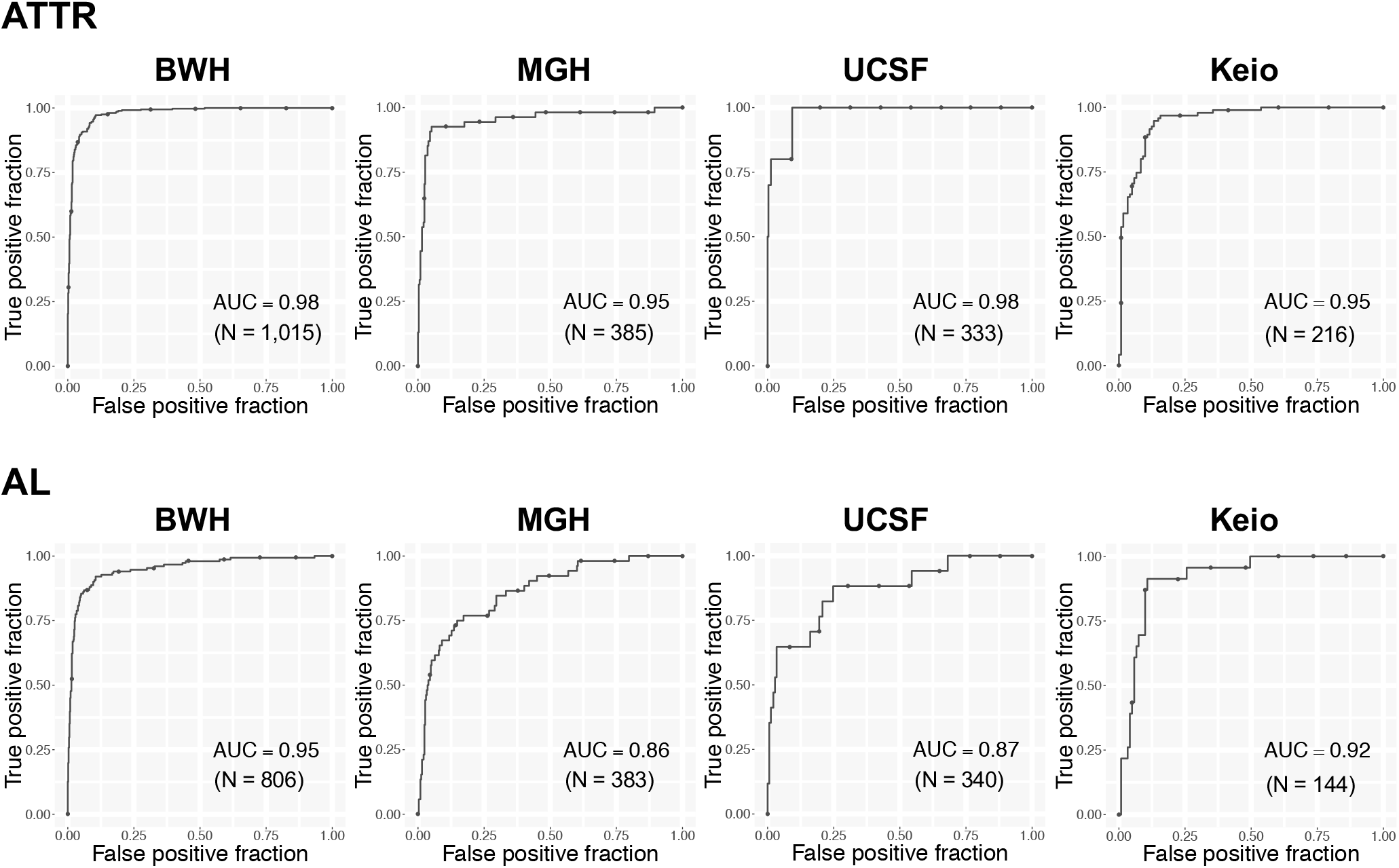
Performance of echocardiogram model on amyloid subtype. ROC plots for detecting cardiac amyloidosis by subtype for each institution. The performance on a test dataset is shown for BWH. N is the numbers of studies

**Figure S3:**
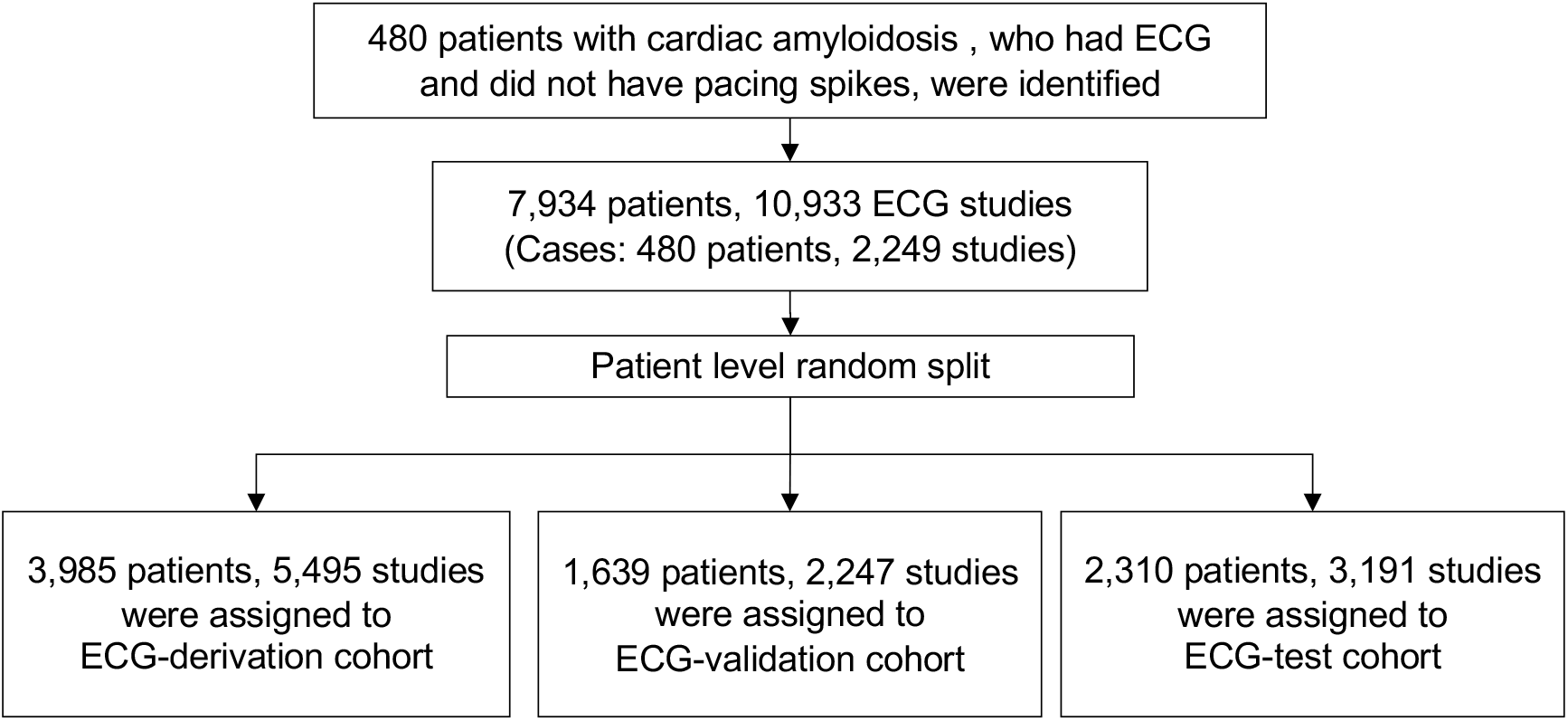
Cohort selection procedure for the BWH ECG cohort.

**Figure S4:**
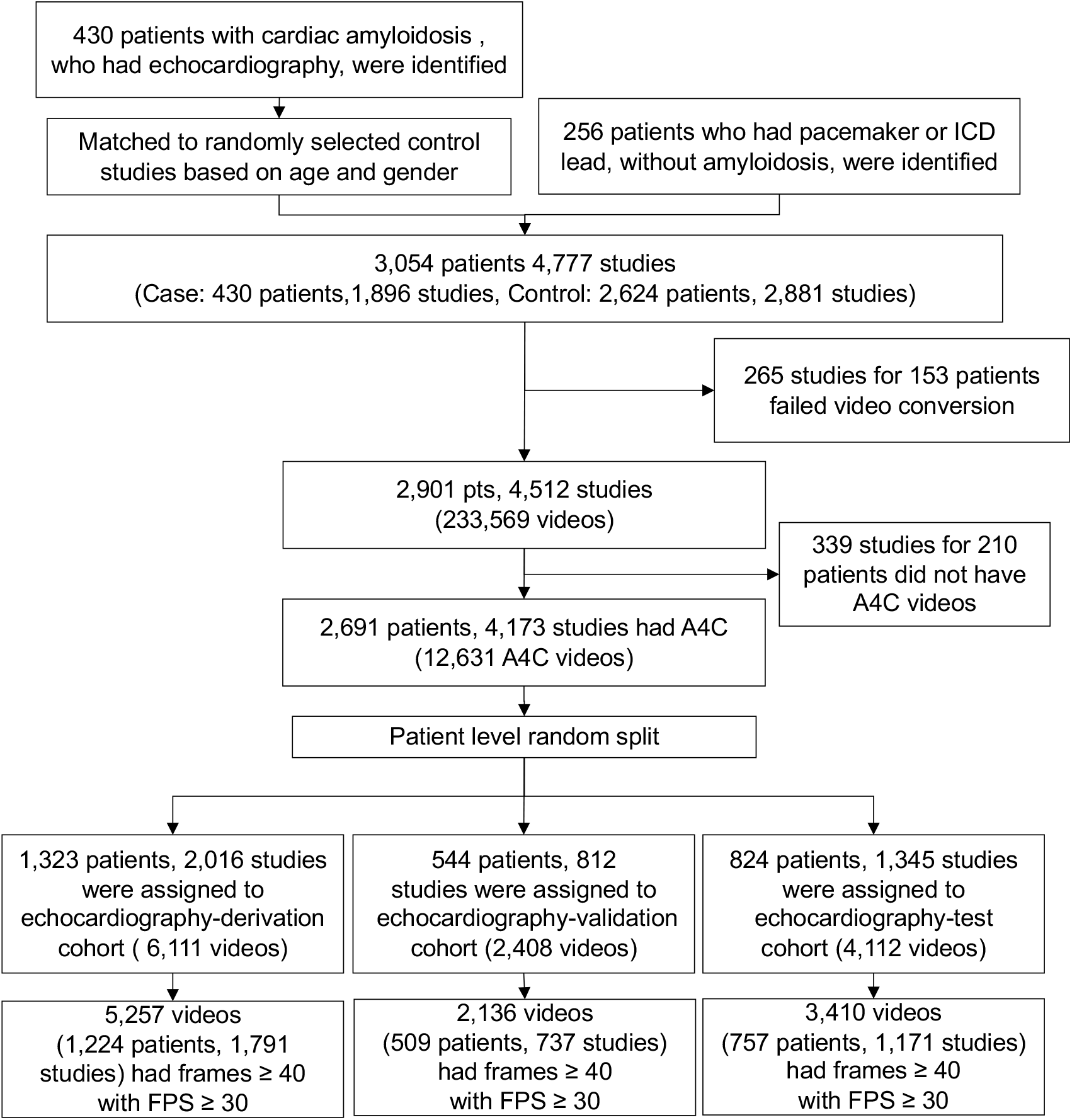
Cohort selection procedure for the BWH echocardiogram cohort.

**Figure S5:**
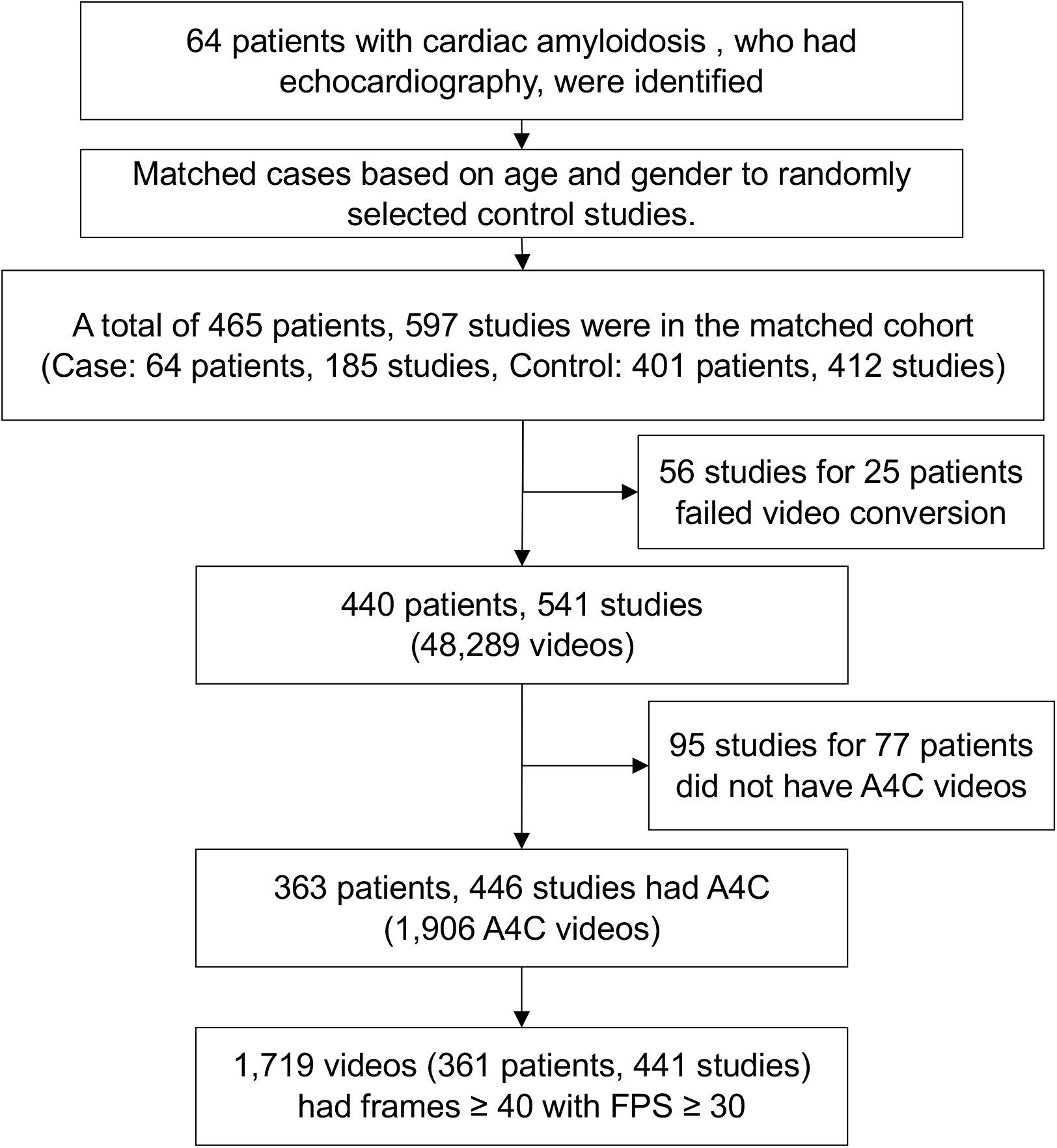
Cohort selection procedure for the MGH echocardiogram cohort.

**Figure S6:**
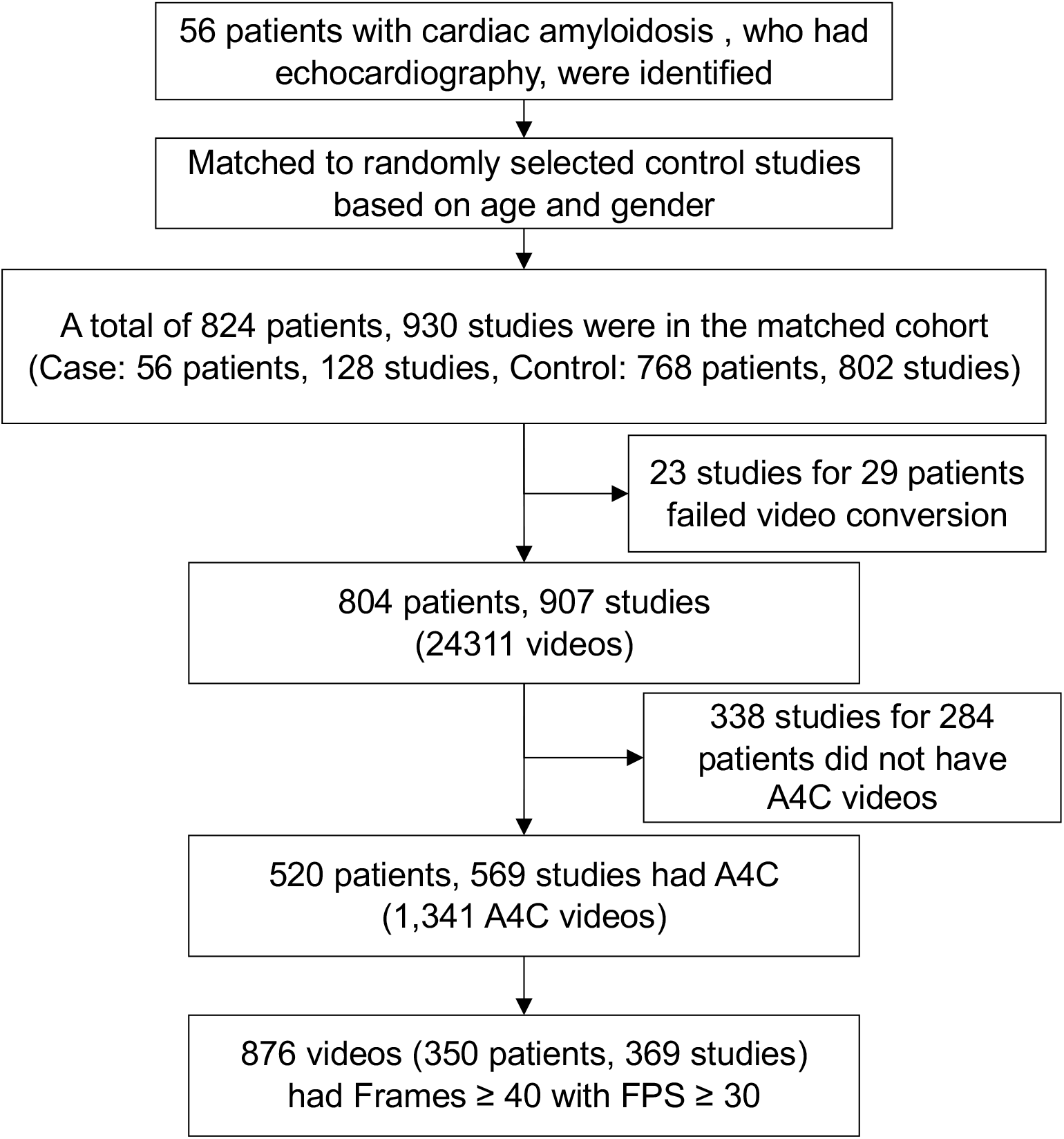
Cohort selection procedure for the UCSF echocardiogram cohort.

**Figure S7:**
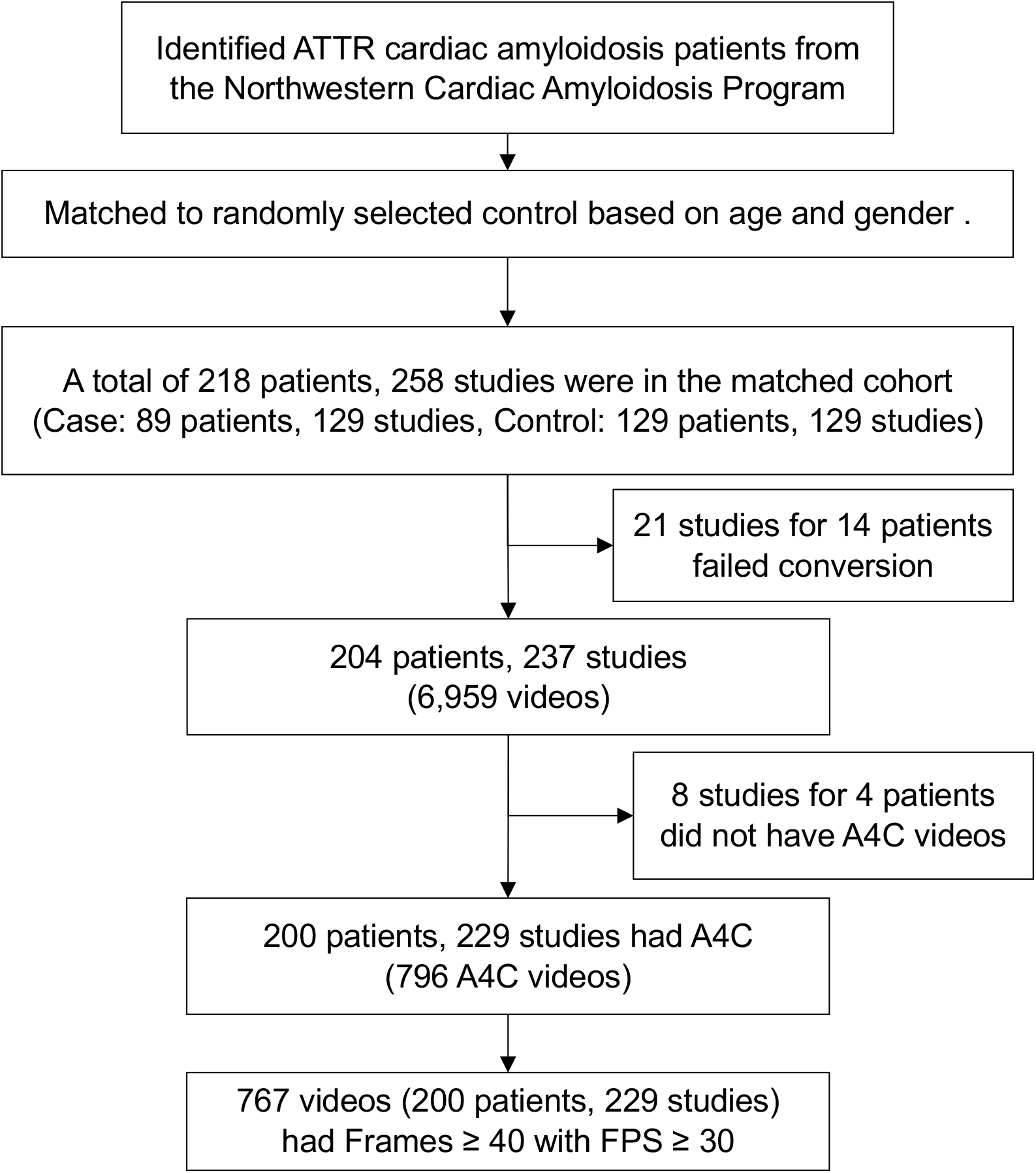
Cohort selection procedure for the NW echocardiogram cohort.

**Figure S8:**
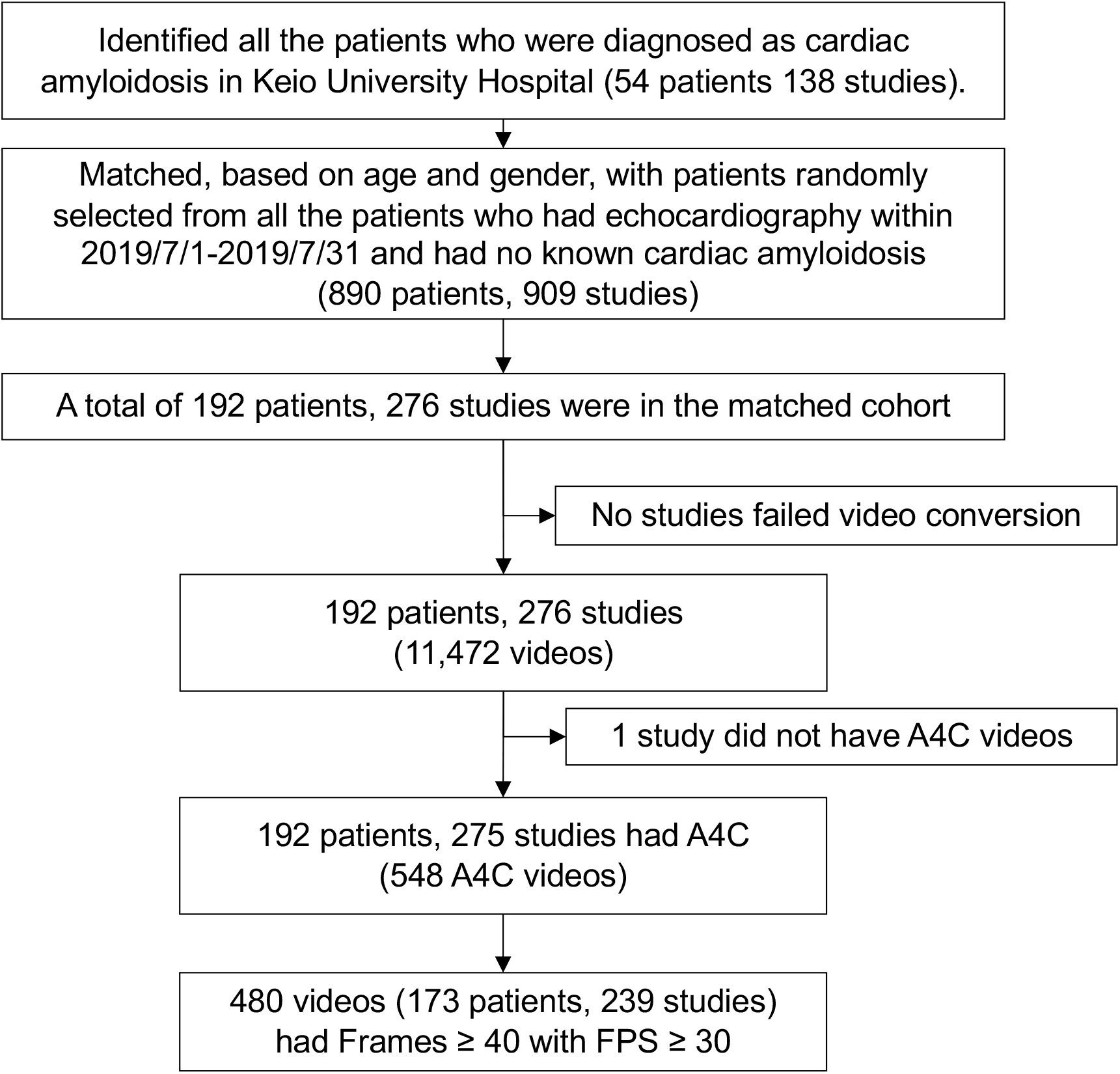
Cohort selection procedure for the Keio echocardiogram cohort.

**Figure S9:**
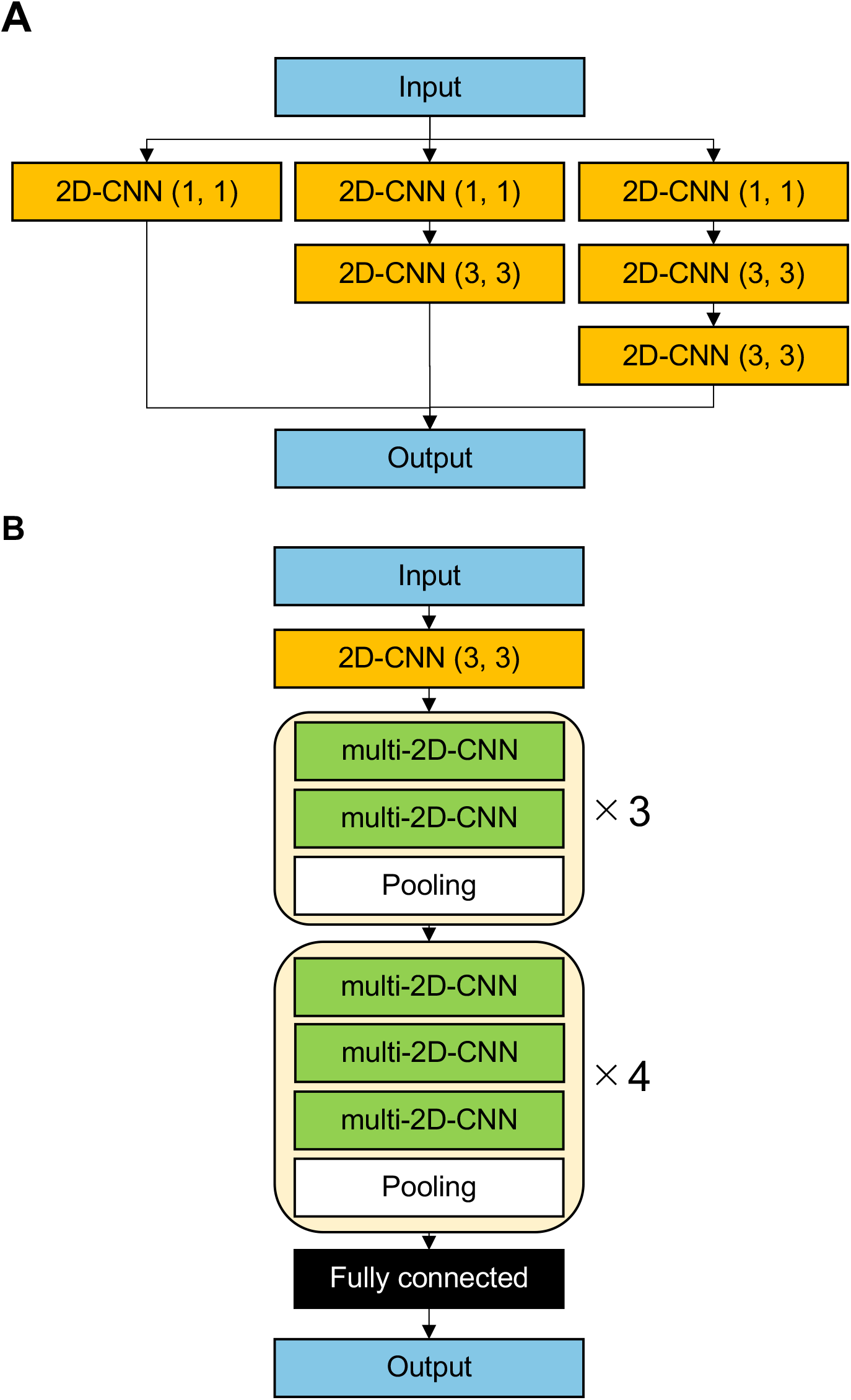
Schematic drawing of the network structure of ECG model. (A) Structure of the multi-2D-CNN module and (B) the full network structure of ECG model.

**Figure S10:**
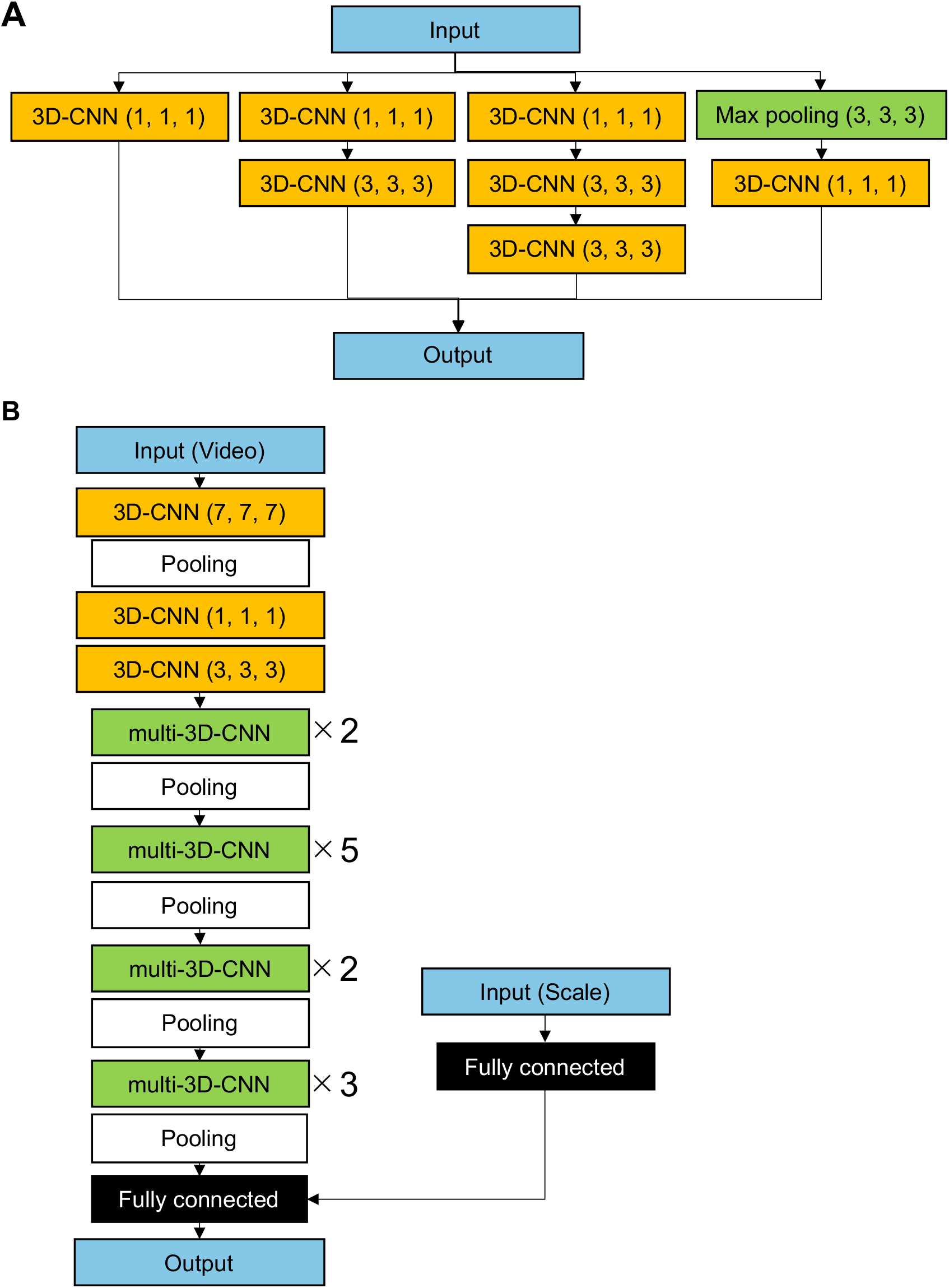
Schematic drawing of the network structure of echo model. (A) Structure of the multi-3D-CNN module and (B) the full network structure of echo model.

**Figure S11:**
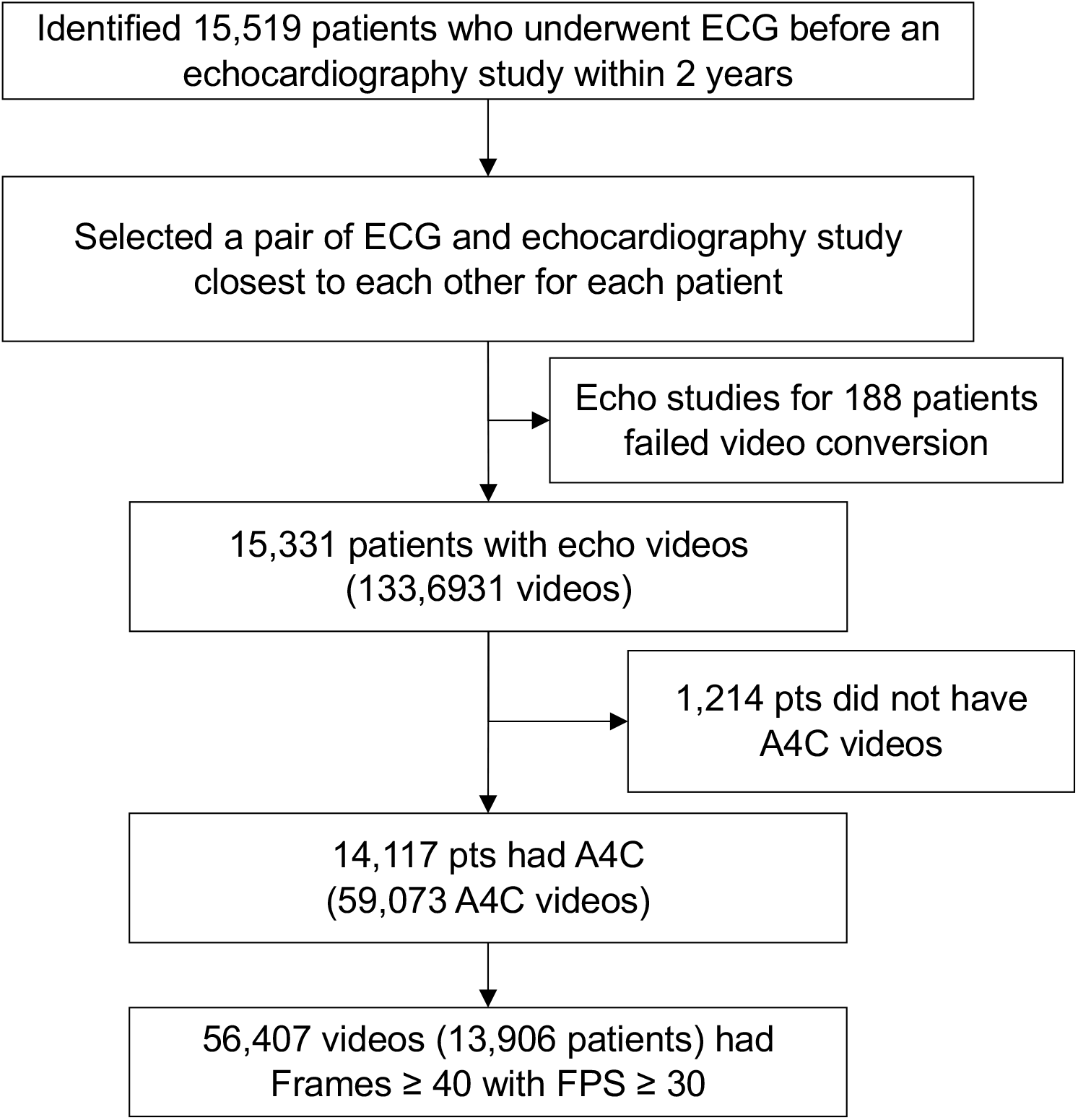
Cohort selection procedure for the MGH surveillance cohort.

**Figure S12:**
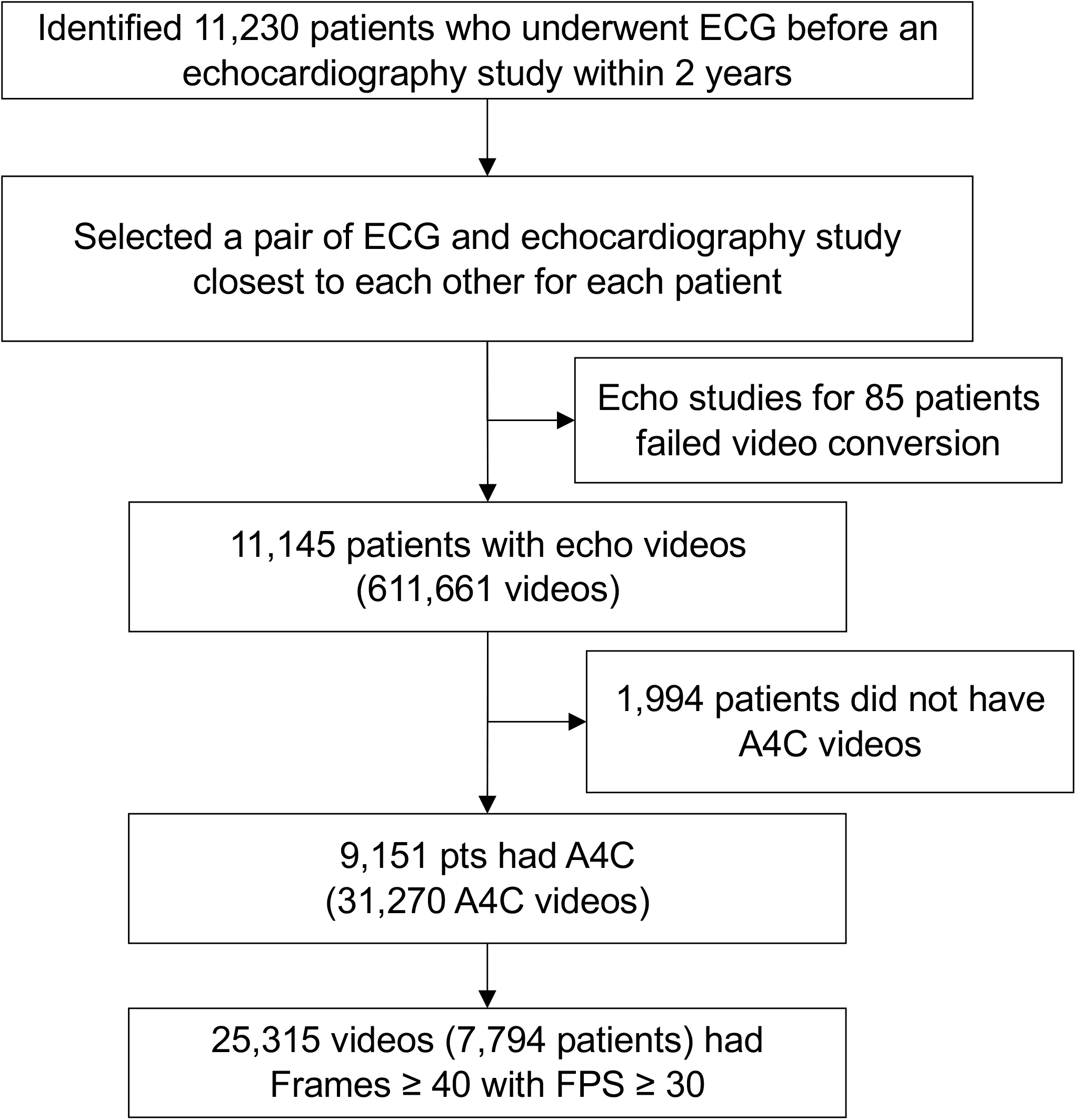
Cohort selection procedure for the MGH surveillance cohort.

